# Heightened Risk of Myocardial Ischemia with Mental Stress Among Black Women Survivors of a Myocardial Infarction in Midlife

**DOI:** 10.1101/2025.06.27.25330451

**Authors:** Viola Vaccarino, Amit J. Shah, Tené T. Lewis, Marina Piccinelli, Lisa Elon, Hua She, Yi-An Ko, Zachary T. Martin, Nancy Murrah, Lucy Shallenberger, Tatum Roberts, Lewam Stefanos, J. Douglas Bremner, Paolo Raggi, Arshed Quyyumi

## Abstract

**Background:** Stark disparities in the outcome of myocardial infarction (MI) persist, with large unexplained variations affecting younger Black women. Mental stress induced myocardial ischemia (MSIMI) is an emerging mechanistic pathway that may help explain excess risks in this group.

**Objectives:** To determine whether MSIMI is more common in Black women with recent premature MI than other demographic groups.

**Methods:** We studied 602 individuals ≤ 61 years of age who were hospitalized for MI in the previous 8 months. Participants underwent 99mTc-sestamibi myocardial perfusion imaging at rest and after mental stress (speech task). A summed difference score (SDS) was used to quantify ischemia. Clinically significant MSIMI was defined as an SDS ≥3. Log-binomial regression models adjusted for sociodemographic, lifestyle, clinical and psychosocial factors.

**Results:** The mean age was 51 years (range, 25-61), 46% were women and 59% self-identified as Black. Black women had a more adverse psychosocial profile and higher rates of obesity and diabetes, but a less severe index MI. The incidence of MSIMI was approximately doubled in Black women than the other groups (p<.001 for interaction). Clinical and psychosocial risk factors did not explain these differences. In a fully adjusted model, the risk ratio of MSIMI for Black women was 2.2 (95% CI, 2.0-2.5) compared to Black men, 2.3 (1.8-2.9) compared to non-Black women, and 1.8 (1.4-2.2) compared to non-Black men.

**Conclusions:** Among midlife individuals with a recent MI, Black women have a disproportionately higher risk of MSIMI. Targeted interventions for this high-risk group are needed.

**CLINICAL PERSPECTIVE:** *What is new?:* We show for the first time that Black women in midlife who have recently experienced a myocardial infarction face a disproportionately high risk of developing myocardial ischemia when under mental stress. This elevated risk cannot be attributed to more severe coronary artery disease, suggesting a stress-related cardiovascular vulnerability that may help explain why Black women experience both higher rates of premature heart attacks and worse outcomes following these events.

*What are the clinical implications?:* The high rate of ischemia with mental stress in Black women highlights the need for new targeted risk assessment protocols and prevention strategies that go beyond the control of conventional risk factors to address stress-related risk pathways. This paradigm shift in cardiac care should help reduce cardiovascular disparities and improve outcomes in this historically underserved population.

**GRAPHICAL ABSTRACT:** *Differences in Myocardial Ischemia with Mental Stress by Sex and Race:* Among 602 individuals ≤ 61 years of age who were hospitalized for a myocardial infarction in the previous 8 months, and who underwent myocardial perfusion imaging with mental stress, Black women had approximately a twofold higher risk of developing myocardial ischemia compared with other demographic groups, even after adjusting for clinical and psychosocial factors. 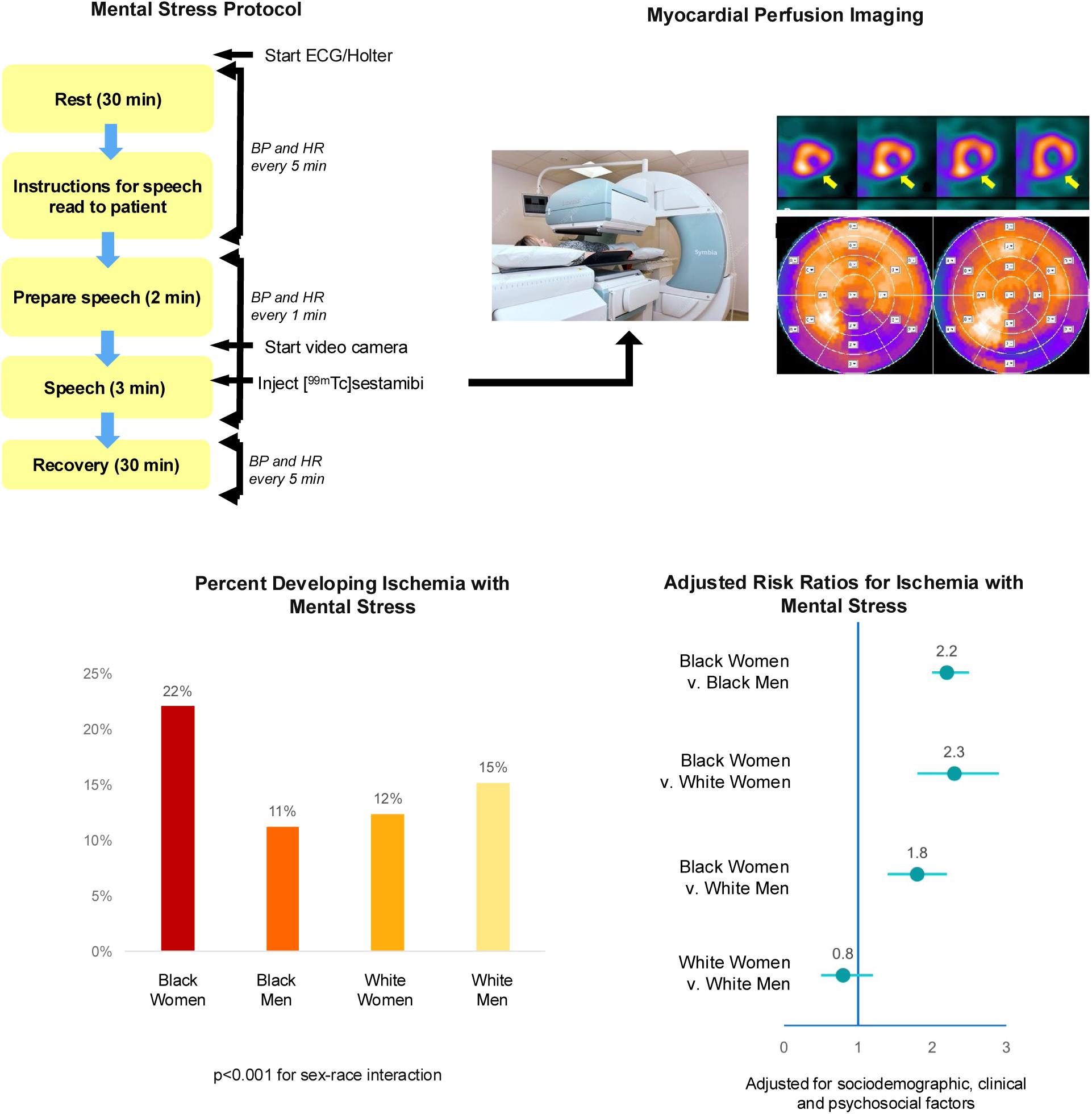

---

Each year, approximately 80,000 women under the age of 65 years are hospitalized for myocardial infarction (MI) in the United States.^1^ These cases constitute roughly 10% of all MI hospitalizations and 30% of MI events occurring before age 65.^1^ Notably, while the incidence of premature MI, typically defined as an MI occurring before age 65 years in women and before age 55 years in men, has remained stable or has slightly declined in men over the past two decades, it has shown a concerning upward trend among women.^2,3^ Thus, premature MI in women represents a growing public health concern.

Black women die from MI at significantly younger ages compared to other demographic groups. Of all MI-related deaths among Black women, 24% are premature, a proportion that is considerably higher than for Black men (18%), White women (13%), and White men (11%).^2^ Even among those who survive the initial MI event, there are stark disparities disadvantaging younger Black women.

Following a first MI between ages 45 and 65 years, Black women face a 28% mortality rate within five years—approximately double the mortality rate of White women and of men of either racial group.^4^ The risk of MI recurrence is also elevated, as Black women under the age of 60 years who have survived an initial MI face approximately three times the likelihood of experiencing a subsequent MI compared to their White counterparts.^5^

Psychosocial stress is increasingly recognized as a significant contributor to cardiovascular risk in women,^6^ but current management strategies continue to focus on traditional risk factors and pharmacological therapies. Black women are especially impacted by stress as they face social disadvantage and discrimination both for sex and for race.^7,8^ Maladaptive coping mechanisms may further elevate their cardiovascular risk.^9^ Among middle-aged Black women from the community, specific stressors—racial discrimination, disproportionate financial responsibilities, and stressful life events affecting them or their social networks—have been linked to cardiometabolic risk factors, subclinical markers of cardiovascular disease (CVD), and clinical CVD.^10–13^ These heightened risks likely extend to Black women who have survived an initial MI. Stress is a stronger risk factor for adverse cardiovascular outcomes in individuals with pre-existing coronary heart disease than in low-risk populations,^14,15^ potentially exacerbating the risk of adverse outcomes for Black women with a premature MI.

Mental stress-induced myocardial ischemia (MSIMI) represents a significant downstream risk pathway connecting stressful exposures with adverse outcomes in people with coronary heart disease. This phenomenon, which is potentially treatable,^16^ is associated with a doubling of recurrent events and mortality—a risk magnitude exceeding that of ischemia provoked by conventional stress testing.^17^ Unlike conventional stress-induced ischemia, MSIMI is believed to reflect autonomic responses to emotional stress on the coronary microvasculature rather than the underlying severity of coronary atherosclerosis. Previous research has documented a preponderance of MSIMI among younger women with coronary heart disease,^18–20^ but whether this sex difference is driven by Black women remains unexamined. This knowledge gap is particularly concerning given the disparities in cardiovascular outcomes involving Black women and the fact that stress-related pathways are potentially addressable.

The current study aimed to investigate whether young and middle-aged Black women with a recent MI experience higher rates of MSIMI compared to other demographic groups, and to identify potential explanatory factors for any observed differences.

## METHODS

### Overview of Study Protocol

This study is based on data collected in the second and third waves of the Myocardial Infarction and Mental Stress study (MIMS).^19^ After an initial pilot study (Wave 1),^21^ participants were enrolled in Wave 2 and Wave 3 following similar methodology. Enrollment spanned from April 2012 to March 2017 for Wave 2 (n=313) and from February 2020 to December 2023 for Wave 3 (n=306).

Participants were recruited from a network of Emory University-affiliated hospitals throughout metropolitan Atlanta. Eligibility criteria included documented hospitalization for type 1 MI within the previous 8 months and age between 18-60 years at the time of MI occurrence. Type 1 MI diagnosis was confirmed through medical record review using standard criteria of elevated troponin levels accompanied by ischemic symptoms and/or electrocardiographic changes or other evidence of myocardial necrosis; the presence of obstructive coronary artery disease (CAD) was not required for inclusion.^22^ Exclusion criteria included severe comorbid medical or psychiatric disorder that could interfere with study results, such as cancer, renal failure, severe uncontrolled hypertension, current alcohol or substance abuse, or schizophrenia; pregnancy or breastfeeding; and current use of immunosuppressant or psychotropic medications other than antidepressants. Potential participants were also excluded if they had unstable angina, acute MI or decompensated heart failure within the week prior to study participation; if they weighed over 450 pounds (due to nuclear imaging equipment limitations); and if study cardiologists deemed it unsafe to discontinue anti-ischemic medications for the required 24-hour period before testing.

Participants underwent a standardized mental stress protocol combined with myocardial perfusion imaging with single-photon emission computed tomography (SPECT) using an established protocol.^23^ All study procedures received approval from the Emory University Institutional Review Board, and written informed consent was obtained from each participant.

### Mental Stress Procedure

Mental stress was induced using a standardized public speaking task following an initial 30-minute rest period.^19,23^ Participants were presented with a scenario with emotional content (a close relative being mistreated in a nursing home) and instructed to develop a realistic narrative around this situation. After a two-minute preparation period, participants delivered a three-minute presentation in front of a video camera and an audience of individuals wearing white coats. Participants were informed that their speech would be evaluated for content, quality, and duration. Blood pressure and heart rate were recorded at five-minute intervals during the resting phase and at one-minute intervals throughout the mental stress task. Subjective distress was assessed using the Subjective Units of Distress Scale,^24^ which quantifies perceived distress on a linear scale from 0 to 100.

### Myocardial Perfusion Imaging

Participants underwent two SPECT myocardial perfusion imaging scans following injection of Technetium-99m ([^99m^Tc])-labeled sestamibi, one at rest and one during mental stress (administered one minute into the speech task). The doses of ^99m^Tc were 10-14 mCi for rest imaging and 30-40 mCi for stress imaging, based on weight. Following standard nuclear cardiology protocols, anti-ischemic medications were withheld for 24 hours prior to testing.

Visual interpretation of the scans was performed by an experienced nuclear cardiologist (PR) who remained blind to demographic and clinical data. The 17-segment model was employed to assess regional myocardial perfusion. Each myocardial segment was scored on a 5-point scale: 0 (normal), 1 (possibly normal), 2 (definitely abnormal), 3 (severely abnormal), and 4 (no perfusion). Standard summed scores were calculated, including a summed stress score (SSS), a summed rest score (SRS), and a summed difference score (SDS).^25^ MSIMI was defined as SDS ≥3.^26^ To confirm robustness of results, additional measures were employed, including the percent ischemic myocardium and the number of ischemic segments.^19,27^ The percent ischemic myocardium, calculated as (SDS/68) × 100, provides a semiquantitative measure of ischemic burden as the percentage of myocardium at risk.

Within individual segments, myocardial ischemia was defined as either a new perfusion abnormality (score ≥2), or worsening of a pre-existing abnormality by ≥2 points in a single segment or ≥1 point in two or more contiguous segments.^17,27^

### Other Measurements

A research nurse obtained detailed sociodemographic, medical history and medication information, and measured weight and height to calculate the BMI. Race was self-reported. Trained personnel abstracted medical records for the index MI including clinical information and angiographic data for severity of CAD prior to any revascularization procedure. Obstructive CAD was defined as ≥ 70% lumen stenosis. Severity of CAD was quantified with the Gensini scoring method, which takes into account degree of luminal stenosis and prognostic significance of coronary tree location.^28^

Validated instruments were employed to assess behavioral, social, and health status information. Depressive symptomatology was evaluated using the Beck Depression Inventory-II (BDI-II).^29^ Psychiatric diagnoses were obtained with the Structured Clinical Interview for DSM-IV (SCID),^30^ which provided lifetime diagnostic information for major depressive disorder and posttraumatic stress disorder (PTSD). The SCID was administered by a research nurse with specialized training, under the supervision of the study psychiatrist (JDB). We also administered the PTSD Checklist for DSM-IV^31^ to assess current PTSD symptoms on a continuous scale,^32^ the Spielberger’s Anxiety Inventory (20-item state anxiety module),^33^ the Spielberger’s Anger Inventory (15-item state anger module),^34^ and the Everyday Discrimination Scale (10-item version) to assess exposure to discrimination or unfair treatment in everyday life.^35^

### Statistical Analysis

Because most of the participants self-identified as either Black/African American or White, we collapsed race categories as Black and Non-Black. However, we repeated the main analyses after excluding individuals of races other than Black or White, to make sure the results remained consistent. We described demographic, behavioral and clinical characteristics and hemodynamic changes during mental stress across four groups based on sex and race. No hypothesis testing was done in this descriptive analysis.^36^

In all models and for all statistical tests we used random effects to preserve the clustering within the two study waves. To assess differences in MSIMI, we fitted log-binomial regression models to directly estimate rate ratios. Using nested models, we sequentially adjusted for sociodemographic and lifestyle characteristics, clinical risk variables, and, lastly, psychosocial factors. In all models we tested interactions by sex and race.

Missing data were minimal; angiographic data had the most missing values (4%). Even so, we performed multiple imputations for the final model to explore the impact of missing data on the results. Data were imputed 50 times using the Markov chain Monte Carlo method of multiple imputation, implemented using SAS PROC MI, and imputed regression estimates were combined via SAS PROC MIANYLYZE.^37^ We also assessed the missing at random (MAR) assumption by differential weighting of variable values (SAS MNAR statement) and found results to be nearly identical, thus justifying reliance on the MAR assumption. The estimates in the imputed analysis matched very closely the not-imputed results, suggesting that bias was minimal or absent. Therefore, we report results based on not-imputed data. All analyses were conducted using SAS statistical software version 9.4 (SAS Inc., Cary, NC).

## RESULTS

### Study Sample

Of the 619 participants in the two waves of the MIMS study, 602 participants (306 in Wave 2 and 296 in Wave 3) had complete imaging results and were included in the present analysis. In the combined sample there were 276 women (46%) and 326 men. The racial composition was predominantly Black/African American (n=356, 59%) or White (202, 34%), with a small number of other races, including 21 Asians, 4 American Indians or Alaska natives, 2 non-Black West Indians, and the remaining were of mixed race or unspecified. Thus, White participants represented 82% of the non-Black group. The sample was primarily middle-aged, with a mean age was 51 years (standard deviation, 6.8 years; range, 25 to 61 years). All participants were younger than 61 years, except for 2 whose birthday occurred after screening but before study visits. The mean and median time from the date of the index MI to the study visit were both 4 months and were similar by sex and race.

### Baseline Characteristics

Both Black women and Black men had a more adverse sociodemographic and psychosocial profile than their non-Black counterparts, including lower income and education and lower likelihood of being married, especially Black women (**Table 1**). The lifetime prevalence of major depression was similarly elevated in Black and non-Black women (41% and 48%, respectively), while it was less than 30% in both male groups. However, psychosocial scale scores for current symptoms of depression and PTSD, as well as scores of state anxiety and state anger tended all to be higher in Black women than in the other groups. Everyday Discrimination Scale scores were higher in Black participants, both women and men, compared with non-Black participants. Black women tended to have higher levels of metabolic risk factors, especially obesity and diabetes mellitus, but the rate of smoking was similar across groups. Medication use was also similar, except for statins and aspirin that were most commonly used by non-Black men, and antidepressants, which were more often used by women compared to men.

**Table 1.** Characteristics of the study population by sex and race.

|  | **Black**<br>**Women**<br>**N=195** | **Black Men**<br>**N=161** | **Non-Black**<br>**Women**<br>**N=81** | **Non-Black**<br>**Men**<br>**N=165** |
| --- | --- | --- | --- | --- |
| **Demographic Factors** |  |  |  |  |
| Age, years, mean (SD) | 50 (7) | 50 (7) | 52 (6) | 52 (6) |
| Married or living with partner, % | 28 | 39 | 64 | 67 |
| Income < \$25,000, % | 46 | 40 | 25 | 12 |
| Education >12 years | 61 | 49 | 70 | 75 |
| Premenopausal, % | 33 | -- | 21 | -- |
| **Psychosocial Risk Factors** |  |  |  |  |
| Lifetime history of major depression, % | 41 | 26 | 48 | 28 |
| Beck Depression Inventory, mean (SD) | 13 (11) | 11 (10) | 11 (10) | 9 (8) |
| Lifetime history of PTSD, % | 22 | 18 | 19 | 8 |
| PTSD Symptom Checklist, mean (SD) | 34 (14) | 30 (14) | 30 (12) | 28 (12) |
| State Anxiety, mean (SD) | 37 (13) | 35 (13) | 29 (9) | 31 (10) |
| State Anger, mean (SD) | 39 (11) | 37 (13) | 31 (9) | 32 (10) |
| Everyday Discrimination Scale, mean (SD) | 18 (6) | 18 (7) | 15 (5) | 16 (5) |
| **Cardiovascular Risk Factors** |  |  |  |  |
| BMI, kg/m<sup>2</sup>, mean (SD) | 33 (7) | 30 (5) | 30 (8) | 30 (5) |
| Ever smoker, % | 49 | 54 | 48 | 42 |
| History of hypertension, % | 88 | 84 | 69 | 69 |
| History of dyslipidemia, % | 76 | 78 | 69 | 83 |
| History of diabetes, % | 41 | 30 | 30 | 28 |
| **Medications** |  |  |  |  |
| Beta Blocker, % | 85 | 87 | 84 | 86 |
| Statins, % | 85 | 86 | 80 | 92 |
| Aspirin, % | 82 | 79 | 88 | 90 |
| ACE Inhibitors, % | 34 | 43 | 29 | 40 |
| ARB medications, % | 28 | 18 | 19 | 18 |
| Antidepressants, % | 21 | 6 | 28 | 14 |
| **Clinical Characteristics** |  |  |  |  |
| ST-segment elevation MI, % | 22 | 31 | 25 | 40 |
| Maximum troponin, ng/mL, mean (SD) | 10 (17) | 13 (20) | 10 (18) | 20 (25) |
| LV ejection fraction (%), mean (SD) | 52 (12) | 48 (12) | 54 (9) | 48 (12) |

**Table 1 (Continued)**
|  |  |  |  |  |
| --- | --- | --- | --- | --- |
| Gensini CAD severity score, mean (SD)*† | 19 (5) | 24 (4) | 17 (5) | 40 (3) |
| Obstructive CAD (stenosis $\geq 70\%$), %† | 77 | 85 | 78 | 91 |
| Three-vessel disease (at $\geq 70\%$), %† | 8 | 9 | 6 | 20 |
| History of congestive heart failure, % | 13 | 9 | 2 | 5 |
| History of MI prior to index MI, % | 24 | 21 | 12 | 18 |
| History of revascularization, prior to, or at the index MI, % | 82 | 84 | 83 | 93 |
| **Resting Hemodynamics** |  |  |  |  |
| Systolic blood pressure, mmHg, mean (SD) | 140 (23) | 135 (20) | 128 (19) | 125 (16) |
| Diastolic blood pressure, mmHg, mean (SD) | 86 (13) | 85 (13) | 78 (11) | 79 (10) |
| Heart rate, beat/min, mean (SD) | 66 (10) | 63 (12) | 66 (11) | 62 (12) |

SD: standard deviation; BMI: Body mass index; ACE: angiotensin-converting enzyme; ARB: angiotensin receptor blocker; MI: myocardial infarction; LV: left ventricle; CAD: coronary artery disease.
\* Geometric means.
† Angiographic data available in 578 patients (96%).

Both Black and non-Black women, when compared to men, had evidence for a less severe index MI, including lower peak troponin levels and higher left ventricular ejection fraction; an ST-segment elevation MI was also less common in women (**Table 1**). Indicators of severe obstructive CAD prior to any revascularization were also less prevalent in women of both racial groups compared to men, as evidenced by a lower Gensini score and a lower prevalence of obstructive CAD. Of all groups, non-Black men had the highest prevalence of obstructive CAD and the highest rate of revascularization. A history of heart failure was infrequent, but it was highest in Black women despite their less severe CAD and higher left ventricular ejection fraction. Black women had also the highest levels of resting systolic and diastolic blood pressure, followed by Black men. Heart rate, however, was similarly elevated in both female groups compared to their male counterparts.

The entire sample showed a robust hemodynamic response to mental stress (**Table 2**). After adjusting for resting levels, hemodynamic responses to mental stress were similar across the four groups. Subjective responses to stress were also similar.

**Table 2.** Differences in hemodynamic and subjective responses to mental stress by sex and race.

|  | **Black Women**<br>**N=195** | **Black Men**<br>**N=161** | **Non-Black**<br>**Women**<br>**N=81** | **Non-Black**<br>**Men**<br>**N=165** | P for sex<br>by race<br>interaction<br>* |
| --- | --- | --- | --- | --- | --- |
| **Mental Stress** |  |  |  |  |  |
| <u>Hemodynamic Responses<sup>†</sup></u> |  |  |  |  |  |
| Systolic blood pressure, mm Hg,<br>mean change (95% CI) | 38 (41, 46) | 40 (37, 42) | 46 (42, 49) | 43 (41, 46) | 0.11 |
| Diastolic blood pressure, mm Hg,<br>mean change (95% CI) | 26 (24, 28) | 27 (26, 29) | 30 (27, 32) | 29 (27, 31) | 0.21 |
| Heart rate, beats/min, mean change<br>(95% CI) | 23 (21, 26) | 21 (18, 24) | 30 (26, 33) | 27 (24, 29) | 0.70 |
| <u>Subjective Responses<sup>‡</sup></u> |  |  |  |  |  |
| Subjective Units of Distress Scale,<br>mean change (95% CI) | 33 (27, 39) | 22 (16, 28) | 35 (28, 42) | 29 (23, 35) | 0.28 |

CI: confidence interval.
\* Data are from models that included a random effect for study wave.
<sup>†</sup> Difference between maximum value during stress and minimum value during rest. Differences are adjusted for pretest values.
<sup>‡</sup> Difference between posttest and pretest values: a positive value indicates higher distress with mental stress. Differences are adjusted for pretest values.

### Myocardial Perfusion

Overall, 96 (16%) of the participants developed MSIMI. The rate was similar in Wave 2 (16.3%) and Wave 3 (15.5%). Among Black women, 22% experienced MSIMI, which was about two-fold the rate in the other groups (**Table 3, Central Illustration**). The higher occurrence of MSIMI in Black women was noted irrespective of whether ischemia was analyzed as a dichotomous variable (presence/absence of ischemia), as a quantitative score (percent of ischemic myocardium), or as a count of ischemic myocardial segments (**Table 3**).

**Table 3.** Unadjusted differences in myocardial ischemia measures by sex and race.

|  | **Black Women** | **Black Men** | **Non-Black Women** | **Non-Black Men** | P for sex by race interaction* |
| --- | --- | --- | --- | --- | --- |
| Myocardial ischemia, n (%) | 43 (22) | 18 (11) | 10 (12) | 25 (15) | <0.001 |
| No. of ischemic segments, mean (95% CI) | 0.6 (0.4, 0.9) | 0.3 (0.2, 0.5) | 0.3 (0.1, 0.6) | 0.5 (0.3, 0.7) | 0.03 |
| Percent left ventricle with inducible ischemia, n (%) |  |  |  |  |  |
| 0% | 139 (71) | 132 (82) | 68 (84) | 126 (76) | <0.001 |
| >0% - <5% | 29 (15) | 19 (12) | 7 (9) | 20 (12) |  |
| ≥5% | 27 (14) | 10 (6) | 6 (7) | 19 (11) |  |

CI: confidence interval.
\* p values are from models that included random effects for study wave.

Adjustment for sociodemographic and lifestyle factors, CAD severity, and psychosocial factors did not materially affect differences in MSIMI by race and sex (**Table 4**). In each of the models, Black women had about twofold higher rate of MSIMI than Black men and non-Black women, and between 40% to 70% higher rate of MSIMI than non-Black men. In contrast, there was no difference in the rate of MSIMI between non-Black women and non-Black men. The p-value for the interaction by sex and race was significant in all models.

**Table 4.** Differences in myocardial ischemia with mental stress by sex and race.

|  | **Black Women**<br>**vs. Black Men** | **Black Women**<br>**vs. White**<br>**Women** | **Black Women**<br>**vs. White Men** | **White Women**<br>**vs. White Men** |  |
| --- | --- | --- | --- | --- | --- |
|  | Risk Ratio (95% CI) |  |  |  | P for sex by<br>race<br>interaction |
| Unadjusted | 2.0 (1.5-2.7) | 1.7 (1.6-1.8) | 1.4 (0.9-2.1) | 0.8 (0.5-1.3) | <0.001 |
| Adjusted for<br>sociodemographic and<br>lifestyle risk factors* | 2.0 (1.7-2.2) | 2.0 (1.7, 2.3) | 1.6 (1.1-2.5) | 0.8 (0.5-1.5) | <0.001 |
| Adjusted for the above plus<br>CAD risk factors and CAD<br>severity† | 2.3 (1.9-2.6) | 2.2 (2.0-2.3) | 2.0 (1.3-3.1) | 0.9 (0.6-1.5) | <0.001 |
| Adjusted for the above plus<br>psychosocial factors‡ and<br>antidepressant use | 2.2 (2.0-2.5) | 2.3 (1.8-2.9) | 1.8 (1.4-2.2) | 0.8 (0.5-1.2) | <0.001 |

CI: confidence interval; CAD: coronary artery disease.
\*Age, race (black versus non-black), married, education >12 years, ever smoking, and BMI.
† Hypertension, diabetes mellitus, left ventricular ejection fraction, Gensini CAD severity score, ST-segment elevation MI, and resting perfusion defect severity (SPECT summed rest score).
‡ Beck Depression Inventory score, PTSD Symptom Checklist score, and Everyday Discrimination Scale score. Substituting depression and PTSD symptom scores with DSM-IV diagnoses for depression and PTSD did not materially change the results.

When the analyses were repeated after excluding participants who self-identified as belonging to race/ethnicity other than Black or White, reducing the sample to 558 participants, the results remained similar. After multiple imputations for missing values, the results also were consistent.

## DISCUSSION

We found that the likelihood of developing MSIMI in middle-aged Black women with a recent MI was double that of non-Black women and of men of either racial group. This was observed even though women, Black and non-Black, had less obstructive CAD than men.

This study represents the first investigation examining differences in MSIMI across both sex and race simultaneously. In previous research based on smaller samples of young and middle-aged post-MI patients, including Wave 1 and 2 of MIMS,^19,21^ we reported higher rates of MSIMI in women compared to men. Similarly, in patients with broadly defined stable CAD, we identified a sex-by-age interaction suggesting that vulnerability to MSIMI is specific to younger women.^18^ However, these earlier studies included too few Black women to analyze this demographic group separately. Other research examining sex differences in MSIMI has yielded mixed results, though these studies predominantly enrolled older and White patients.^38,39^

Despite a thorough set of measurements spanning psychosocial and clinical variables, we were unable to explain the excess risk of MSIMI in Black women. Middle-aged Black women in the U.S. face stark health disparities compared to other groups of corresponding age, including higher rates of CVD, hypertension and obesity, and adverse outcomes after an MI.^2,4^ For these women, psychosocial stress can manifest through discrimination, which encompasses harassment, prejudice, and race-sex stereotyping.^9^ Their limited access to resources to counterbalance adversities can lead to socioculturally specific coping strategies, such as the “superwoman schema,” characterized by attempts to maintain resilience despite social adversity.^9^ This coping style has been related to abnormal vascular function in Black women.^40,41^ Discrimination and other social stressors are related to cardiometabolic risk and subclinical and clinical CVD among younger Black women.^10–12^ These elevated risks likely extend to Black women with coronary heart disease. Of note, everyday discrimination was previously related to MSIMI in post-MI populations—and the association was particularly pronounced among Black women.^42^ In our sample, however, everyday discrimination was reported at a similar level in Black women and Black men, and including this factor in the model did not explain the excess risk of MSIMI among Black women. As we did not have information on other sociocultural factors that may be especially relevant for Black women, these should be the subject of future investigation.

Evidence suggests that MSIMI has a microvascular component.^20,43^ Since CAD severity did not explain the higher rates of MSIMI among Black women in our study, microvascular mechanisms may play a role in this disparity, which can help guide the management of this at-risk group. Of note, a decrease in microvascular function during mental stress has been linked to adverse cardiovascular outcomes in women but not in men.^44^ Despite growing data on stress response, microvascular function and MSIMI in women generally,^6^ specific data for Black women remain limited. Collectively, our data suggest that microcirculatory dysfunction and its role in MSIMI may be particularly relevant for understanding premature cardiovascular risk in Black women.

Our findings have substantial implications. The relative proportion of premature MI is escalating among women, and especially among Black women.^2^ Despite considerable progress in improving the awareness of cardiovascular disease as a major cause of morbidity and mortality in women, such awareness has not improved, particularly among minority and younger women.^45,46^ Inadequate recognition of alternative, unmeasured risk factors relevant to the cardiovascular health of Black women may undermine both primary and secondary prevention efforts for this group.^47,48^ The high rate of MSIMI with mental stress we observed in Black women, combined with their adverse psychosocial profile, necessitates a renewed focus on stress-related risk pathways for this vulnerable population. The routine management of CAD patients mostly focuses on controlling conventional cardiovascular risk factors, while psychological aspects remain minimally appreciated.^49^ Our findings highlight the need for targeted risk assessment protocols and prevention strategies specifically designed to address the unique risks of this group.

Our study has limitations. We lacked outcome data, and therefore the clinical significance of our findings needs further study. While the prognostic significance of MSIMI has been established,^17^ more data are needed specifically for Black patients. The speech task for mental stress testing may provide differential responses by sex and race. However, this is an established mental stress protocol when using [^99m^Tc]-based perfusion imaging; the radioisotope injection during mental stress captures a precise “snapshot” of myocardial perfusion during stress. We cannot exclude selection bias due to potential exclusion of those who were too sick or otherwise unable to participate; this may have led to underestimation of the true effect size. As our study was conducted at a single institution, the generalizability of our findings to other populations and settings requires further exploration. Despite its limitations, this represents the only study to date of MSIMI in middle-aged Black women after MI.

In conclusion, among midlife patients with a recent MI, Black women have a disproportionate propensity for myocardial ischemia with mental stress. These findings suggest that cardiovascular responses to psychological stress are implicated in the premature cardiovascular risk and mortality of Black women. Integrating psychological and behavioral factors into secondary prevention represents a critical research priority, as such approaches could help reduce sex and race disparities in post-MI outcomes.

## ABBREVIATIONS LIST

CAD: coronary artery disease
CVD: cardiovascular disease
MI: myocardial Infarction
MSIMI: mental stress-induced myocardial ischemia
MIMS: Myocardial Infarction and Mental Stress Study

## Data Availability

Data will made available upon reasonable request.

